# A scoping review of the research integrity architecture and how it is addressed in legal frameworks, institutional policies, and the scholarly literature: Research protocol

**DOI:** 10.1101/2021.10.15.21265065

**Authors:** Juan Guillermo Pérez, Carolina Torres-Sarmiento, Andrés Felipe Duarte Castro, Luis Eduardo Gómez, Vivienne C. Bachelet

**Author notes:** Correspondence author: Juan Guillermo Pérez.

## Abstract

**Background:** Research integrity is a dynamic area within the ethical research ecosystem. Several efforts have been made to incorporate this topic in scientific governance frameworks. However, the efforts generally result in non-binding declarations and policies. Due to differences in legal systems, research cultures, and institutional approaches worldwide, there is a need to identify and map existent strategies on sound scientific practices.

**Objective:** This scoping review aims to systematically search, map, and evaluate the best available evidence on strategies and recommendations regarding research integrity. The goal is to identify international, national, regional, and local legal frameworks, institutional policies and guidelines, research integrity policies, interventions, strategies, and recommendations for:

i. The design and conduct of research projects,
ii. The publication of research results,
iii. The monitoring of scientific practices,
iv. The implementation of corrective actions, and
v. Mentoring and education on research integrity.

**Methods:** The search will follow the PRISMA Extension for Scoping Reviews (PRISMA-ScR) and the methodological approach designed by Arksey and O’Malley. It will include legal frameworks, national and international governmental and non-governmental documentation, and scholarly articles published in peer-reviewed journals on research integrity. The search will be conducted in PubMed/MEDLINE, Web of Science, JSTOR, Latin American & Caribbean Health Sciences Literature (Lilacs), Scopus, OECD Library. It will be complemented with hand searching and scanning, covering other databases and grey literature sources. We will extract and synthesize the data using two macro-genres: legal documents (soft law and hard law) and non-legal documents.

## 1 Rationale

Research integrity is a dynamic area recognized as vitally important by multiple stakeholders, including governments, funding institutions and the global scientific community ^1,2^. Recently, the topic has been increasingly under the spotlight because of the necessity of creating appropriate scientific governance and other efforts to formalize and institutionalize good science ^3^. According to Armond et al., academic interest in research integrity surged in the last decade for reasons such as the evolving nature of research environments due to the introduction of new technologies, the pressure to publish, competition for funding, diversification in collaboration, and the rise in publicized cases of misconduct ^4^.

Due to its heterogeneity, there is no international consensus about the definition of research integrity ^1^. Different terminologies have been used, such as ‘scientific integrity’, ‘responsible conduct of research’, and ‘research integrity’ ^5^, potentially resulting in ambiguity. Ultimately, it is often up to researchers, institutions, and other external and internal players to come up with definitions ^6^. For investigators, research integrity is related to principles such as honesty, accountability, professional courtesy and fairness, and good stewardship ^7^. For institutions, research integrity may be associated with creating and sustaining environments that promote responsible behaviors and high ethical standards, education, and policies ^7^.

Several efforts have been made internationally to create a roadmap that links research integrity with principles, responsible behaviors, and good practices. For instance, the Singapore Statement on Research Integrity incorporated some principles and professional responsibilities, recognizing the existing and potential national and disciplinary dissimilarities in designing and conducting research ^7^.

Also, the Montreal Statement on Research Integrity in Cross-Boundary Research Collaborations highlighted that cross-national, institutional, disciplinary and sectoral research collaborations are crucial to advancing knowledge ^8^. Specifically, it recognized that these collaborations are particularly challenging for the responsible conduct of research as they potentially encompass significant differences “in regulatory and legal systems, organizational and funding structures, research cultures, and approaches to training” ^8^.

The Hong Kong Principles for Assessing Researchers explicitly focused on strengthening research integrity by rewarding behaviors related to responsible research practices, thus, avoiding “detrimental research practices” ^9^. Moreover, the Hong Kong principles were designed to help institutions “minimize perverse incentives that invite to engage in questionable research practices”^9^.

Furthermore, the Universal Declaration on Bioethics and Human Rights embodies an important step in research integrity by acknowledging that unethical scientific and technological conduct has a distinct impact on peoples and local communities. This declaration focuses on the special needs of developing countries and promotes equitable access to science and technology and the rapid sharing of knowledge ^10^.

In Latin America, countries such as Argentina, Brazil, Chile, Colombia, Costa Rica, Mexico, Nicaragua, Peru, and Venezuela adopted laws, policies and guidelines on research integrity ^11^. However, these frameworks contain limitations such as the lack of agreed definitions, problems with reporting scientific misconduct, deficiencies in the design and implementation of legal standards, absence of regulatory agencies and infrastructure, lack of funding, confusion about roles and responsibilities, miscommunication, and uncertainty ^11^. These constraints make research integrity more challenging to investigate or to understand as a social problem. While research integrity is more institutionalized in the global north, such as North America and Europe, in Latin America, inter-institutional and intersectoral discussions are lacking ^12^

In Colombia, the Ministry of Science, Technology, and Innovation (MinCiencias)—previously named the Administrative Department of Science, Technology, and Innovation (Colciencias)—enacted the National Resolution 0314 of 2018 that implemented the Ethics, Bioethics and Research Integrity Policy ^13^. In general terms, the document incorporates minimum guidelines on ethics and good scientific practices for all actors of the National System of Science, Technology, and Innovation (NSSTaI). The document addresses research integrity as the central component to maintain trust and credibility in science through sound knowledge generation and adoption practices by the national research community. The policy encourages the consolidation of a governance system that encompasses research integrity principles, research ethics, and bioethics at the national level ^13^.

This national policy acknowledges (i) the need for the implementation of internal regulations, as these actions are typically managed autonomously; (ii) the lack of consolidated national data on the different practices (or breaches) concerning, for instance, author intellectual property (Law 23 of 1982); (iii) the absence of legal or disciplinary actions in cases of misconduct which reinforces its invisibility; and (iv) the non-existence of a shared culture on research practices and levels of responsibility, either institutional or personal ^14^.

In sum, research integrity is part of the ethical research ecosystem based on public trust. Scientific misconduct curtails the advancement of knowledge and the social backing of science. Consequently, there is a need for identifying and mapping existing strategies on good scientific practices and the best available evidence in the literature for effective implementation of research integrity programs. Even though there are multiple international efforts to consolidate global frameworks on research integrity, the topic has not been studied extensively in Latin America ^12^.

## 2 Objectives

The purpose of this review is to systematically search, map, and evaluate the best available evidence on strategies and recommendations regarding research integrity and good scientific practices. We aim to identify international, national, regional, and local legal frameworks, institutional policies and guidelines, research integrity policies, interventions, strategies, and recommendations for:

i. The design and conduct of research projects,
ii. The publication of research results,
iii. The monitoring of scientific practices in research-based environments,
iv. The implementation of corrective actions when necessary, and
v. Mentoring and education on research integrity.

## 3 Methods

### 3.1 Protocol and registration

To address the purpose of this review, we will use the scoping review methodology following the PRISMA Extension for Scoping Reviews (PRISMA-ScR). We will also refer to the seminal paper by Arksey and O’Malley on a methodological framework for scoping reviews ^15^. This protocol will be published in a preprint server before beginning the data extraction and evidence synthesis. Quality appraisal or risk of bias assessment will not be done as this review aims to map all documents deemed pertinent to the research field.

### 3.2 Eligibility criteria

Our inclusion criteria will be comprehensive to increase the sensitivity of our search strategies. Thus, we will include:

- International, national, regional, and local legal frameworks involving research integrity, including international declarations, statements, rules, regulations, guidelines, policies, country experiences, case reports and white papers;
- National and international governmental and non-governmental documentation on research integrity;
- Scholarly articles published in peer-reviewed journals with no limitation on study design.

We will not restrict for geographical location. We will include multi and interdisciplinary studies using quantitative, qualitative, mixed methods, and policy papers. We will not exclude any specific areas of scientific knowledge. No limits on publication dates will be applied. We will include references in Spanish and English.

We will exclude any document type or scholarly article not directly addressing research integrity. Likewise, we will not include press releases, opinion pieces, news items, blogs, congress abstracts (unless a full-text version is available), videos, slides, interviews, legal cases, letters to the editor, corrigendum and errata, duplicate publications, scientific integrity reviews, and any other non-scholarly literature.

We will work with two macro-genres: legal documents (soft law and hard law) and non-legal documents.

On the one hand, soft law refers to non-legally binding principles, statements, and declarations; on the other hand, hard law refers to documents that are legally binding and enforceable in courts to the parties involved in the agreements (such as citizens, companies, and governmental institutions in a national jurisdiction or countries in an international or multilateral jurisdiction) ^16^. Accordingly, we will consider international, regional, national, and local legal frameworks and international treaties and statements. Except for the OECD iLibrary, the information sources used to find these legal documents will be hand searched. We will consider the scholarly literature, grey literature and discussion and guideline papers issued by scientific associations and non-governmental entities.

### 3.3 Information sources

To achieve a comprehensive understanding of the state-of-the-art in research integrity in the three different contexts that we are interested in (international, regional, and national/local), we will use the following data sources:

- *PubMed/MEDLINE*: developed by the National Library of Medicine, has more than 22 million references of the biomedical literature, including journals and e-books.
- *Web of Science*: a multidisciplinary database with bibliographic information of around 12.000 international journals, including other open access sources.
- *JSTOR*: a digital library encompassing books and other primary sources, journals in humanities and social sciences. It provides full-text searches of approximately 2,000 international journals.
- *Latin American & Caribbean Health Sciences Literature (Lilacs)*: a database maintained and updated by educational, research and, health institutions from the government and private sector.
- *Scopus* is Elsevier’s abstract and citations database: containing more than 16.500 peer-reviewed journals in different fields, including life sciences, social sciences, physical sciences, and health sciences.
- *OECD Library*: the official online library of the Organization for Economic Cooperation and Development and contains books, papers, and statistical resources.
- *Other information sources:*
  - Global governmental and non-governmental sources and databases (e.g., Australian Government-Australian Research Council. Available at: https://www.arc.gov.au/policies-strategies/research-integrity; The UK Research Integrity Office. Available at: https://ukrio.org; The US Department of Health and Human Services-Office of Research Integrity. Available at: https://ori.hhs.gov).
  - *Open Grey*: a European grey literature information system that contains topics in science, technology, biomedical sciences, among others. Available at: http://www.opengrey.eu/
  - Multilateral organization’s sources and databases (e.g., The World Bank, The World Health Organization, United Nations Educational, Scientific and Cultural Organization (UNESCO)).
  - *Community Research and Development Information Service (CORDIS)*: the European Commission’s main source of results from funded research and innovation projects (FP1 to Horizon 2020). Available at: https://cordis.europa.eu/en *The National Academies of Sciences, Engineering, and Medicine*.
  - *The National Academies Press* publishes reports from the National Academies of Sciences, Engineering, and Medicine: It issues more than 200 books a year and provides information on science and health policy matters. Available at: https://www.nap.edu/catalog/21896/fostering-integrity-in-research
  - *Committee on Publication Ethics (COPE) website: COPE provides leadership in thinking on publication ethics and practical resources to educate and support members, and offers a professional voice in current debates*.

### 3.4 Search strategy

Table 1 shows the terms that will be used to build the search strategies for each data source.

**Table 1:** Terms used for search strategies by language.

| Terms in English | Terms in Spanish |
| --- | --- |
| Research integrity | Integridad en la investigación |
| Publication ethics | Ética en las publicaciones |
| Scientific misconduct | Mala práctica, mala conducta científica |
| Scientific integrity | Integridad científica |
| Responsible research practices | Prácticas responsables en investigación |
| Good research practices | Buenas prácticas en investigación |
| Responsible behaviors in research | Conducta responsable en investigación |
| Research integrity training | Formación sobre la integridad en la investigación |
| Supervision and mentoring | Supervisión y tutoría |
| Training | Formación |
| Research collaboration | Colaboración en la investigación |
| Data management practices | Gestión de datos |
| Strategies | Estrategias |
| Policies | Políticas |

We built the search strategies using a combination of the terms included in Table 1 with the Boolean operators “and” and “or.” These operators were commonly used to represent binary logic values between the representation of keywords.

To be consistent with the research question, we connected the terms to the following categories to structure the search strategies: (i) topic, (ii) population, (iii) intervention, and (iv) type of document. In addition, we used the best combination of terms for each database given the differences between search engines, so the search details will have slight variations to achieve breadth and scope.

PubMed/MEDLINE was used for the initial search strategy test. Because of its wide variety of options, we were able to build a more complex search strategy for this database. In contrast, JSTOR sets a limit of seven terms and 200 characters for search details, entailing fewer query terms than other search engines. In other cases, such as the OECD iLibrary, CORDIS, and Lilacs, the results of the research queries with few terms were enough for this scoping review.

Below, we present the resulting search strategies by information source and the results accrued on the day the search was performed for testing purposes. All results will be imported into a collaborative systematic review software.

#### 3.4.1 PUBMED/MEDLINE

|  |
| --- |
| Query |
| ("research integrity"[Title/Abstract] OR "publication ethics"[Title/Abstract] OR "scientific integrity"[Title/Abstract] OR "research collaboration"[Title/Abstract]) AND (strategies[Title/Abstract] OR policies[Title/Abstract] OR guidelines[Title/Abstract] OR "qualitative study"[Title/Abstract] OR "quantitative study"[Title/Abstract] OR "international treaties"[Title/Abstract] OR "international declarations and statements"[Title/Abstract] OR statutes[Title/Abstract] OR laws[Title/Abstract] OR regulations[Title/Abstract] OR resolutions[Title/Abstract] OR codes[Title/Abstract] OR rules[Title/Abstract] OR normativity[Title/Abstract] OR statement[Title/Abstract]) |

#### 3.4.2 WEB OF SCIENCE

|  |
| --- |
| Query |
| (((((ALL=("research integrity")))) AND ALL=("publication ethics") OR ("scientific misconduct")) |

#### 3.4.3 JSTOR

|  |
| --- |
| Query |
| "research integrity" OR "publication ethics" |

#### 3.4.4 LILACS

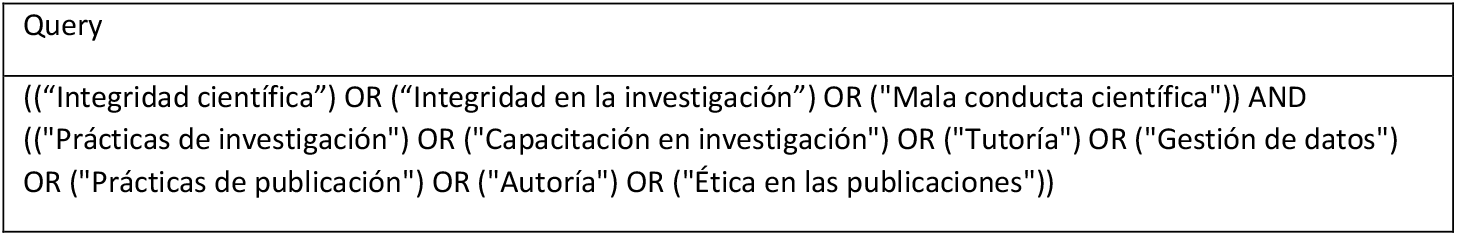

|  |
| --- |
| Query |
| ((("Integridad científica") OR ("Integridad en la investigación") OR ("Mala conducta científica")) AND ((("Prácticas de investigación") OR ("Capacitación en investigación") OR ("Tutoría") OR ("Gestión de datos") OR ("Prácticas de publicación") OR ("Autoría") OR ("Ética en las publicaciones")) |

#### 3.4.5 SCOPUS

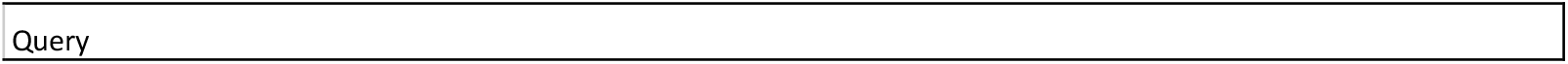
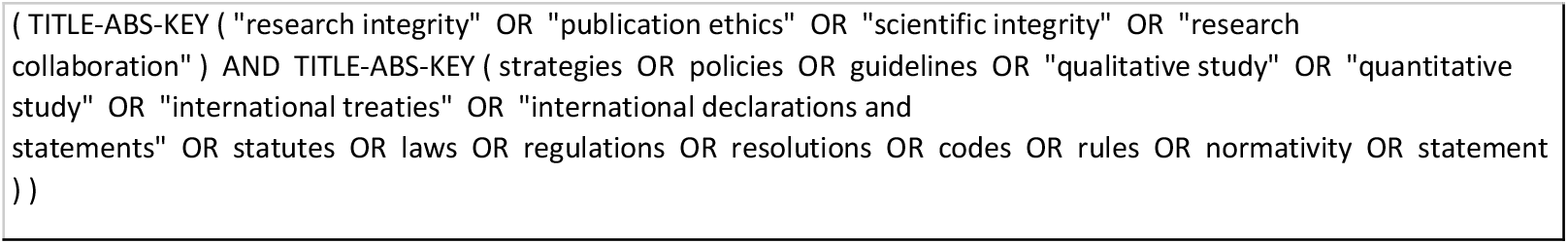

#### 3.4.6 OECD ILibrary

|  |
| --- |
| Query |
| "research integrity" OR "publication ethics" |

#### 3.4.7 Other information sources

For the information sources, we will apply on the search bar the terms related to research integrity shown in Table 1. We expect to find the best available documents on a case-by-case basis, complemented with hand searching and scanning.

### 3.5 Selection process

All retrieved documents—either from databases or through hand searching of websites and additional grey literature sources—will be compiled in Mendeley, a reference management software, where they will be tagged by source provenance. From Mendeley, the identified documents will be imported into Rayyan, collaborative software for systematic reviews, after removing duplicates.

Using Rayyan, four reviewers (CT, LG, FD, JB) working in pairs will independently screen titles and abstracts.

For calibration purposes, each reviewer will screen the same set of ten randomly selected documents to ensure consistent use of the inclusion and exclusion criteria among the evaluators. We will repeat this task as many times as necessary until an 80% concordance is reached between the reviewers.

Once calibration has been achieved, the four evaluators will work in pairs and screen the whole population of identified documents independently and in parallel, thus ensuring an impartial and blinded screening process. Discrepancies will be resolved by consensus with the participation of two senior investigators (VCB and JGP).

When the title and abstract screening process has been completed, the full text of the eligible documents will be retrieved. Again, applying the inclusion and exclusion criteria, the four evaluators working in pairs will select the records for inclusion into this scoping review. As before, discrepancies will be resolved after discussion with a third, more experienced reviewer or with group discussion.

Because the reviewers will become increasingly familiarized with the retrieved documents and more knowledgeable with the search results, to increase the precision of the results, an interim analysis will be done when at least 10% of the documents or a threshold of 50 papers—whichever comes first—have been assessed for inclusion after the full-text evaluation to reassess and adjust the inclusion and exclusion criteria, if need be.

### 3.6 Data charting process

We will create a form using Google Sheets to extract the relevant information from the information sources. Each of the four reviewers will be assigned one-fourth of the included documents and will extract the data items. Another reviewer will cross-check the data extraction to ensure accuracy. The senior investigators will quality check the charting process regularly to achieve consistency of extraction among the reviewers and with the scoping study objectives, and the charting form will iteratively be adapted as needed. No authors will be contacted during the data extraction phase.

### 3.7 Data items

We will extract the following information from the full text of the selected non-legal documents: (I) first author; (II) title; (III) year of publication; (IV) country; (V) article genre (e.g., internal policies, internal guidelines, original research, statements); (VI) key words; (VIII) other participants including institution(s) and person(s) involved; (IX) discipline.

For legal documents, we will use the following categorization criteria: (I) author (multilateral organizations, and states and states agencies); (II) title; (III) type of legal document (treaty, statement, law, policies, guidelines, etc.); (IV) jurisdiction (international, regional, national, and local); (V) year of publication.

Regarding all selected documents, we will extract the following data items for thematic analysis:

- Objective
- Main results and conclusions
- Recommendations

Other additional data items might be incorporated during the data extraction process.

### 3.8 Critical appraisal

As stated previously, no critical appraisal or risk of bias assessment will be done due to the nature of the topic and the need to map all documents that fulfill the inclusion criteria.

### 3.9 Synthesis of results

Using Google Sheet to collect, summarize, and compare the extracted information for each selected document, we will do thematic analysis on the included documents based on Braun and Clarke’s ^17^ thematic analysis methodological approach, which consists of six phases: (i) read and reread the data to become familiar with it; (ii) generate initial nodes; (iii) search for themes; (iv) review the themes; (v) define and name the themes; and (vi) elaborate the report.

The extracted information will provide insight on objectives, main results, conclusions, and recommendations, allowing us to understand the impact that population, strategies, findings, and results have on the design of research integrity normativity, as well as providing an overview of any stakeholders’ role in research integrity development on the legal side. We will also be able to map by jurisdiction, thus helping us identify each legal system’s singularities.

## Data Availability

All data produced in the present study are available upon reasonable request to the authors.

## 4 Notes

### Ethics

This scoping review goes by the research ethics framework of Colombia and the main international statements on research ethics (such as Helsinki and Taipei declarations). Ethics committee approval was granted by Pontificia Universidad Javeriana – Cali (August 26, 2019).

### Author contributions

JGP contributed to conceptualization, funding acquisition, investigation, methodology, project administration, supervision, validation, and review of the original draft. CT, FD, and LG contributed to the conceptualization, data curation, investigation, methodology, and writing of the original draft. VCB contributed to conceptualization, investigation, methodology, project administration, supervision, validation, and review and editing of the original draft.

## Ackowlegments

Special thanks to Jackeline Bravo Chamorro, Carlos Enrique Trillos, and Diana Bernal for their insights and feedback on behalf of the GREICI project investigators.

## 6 Search strategies per data source

Date of search: September 27, 2021.

### 6.1 Search Strategy for PubMed/MEDLINE

|  |  | Query | Hits |
| --- | --- | --- | --- |
| Topic | 1 | "research integrity"[Title/Abstract] OR "publication ethics"[Title/Abstract] OR "scientific integrity"[Title/Abstract] OR "research collaboration"[Title/Abstract] | 3,134 |
| Type of document | 2 | strategies OR policies OR guidelines OR "qualitative study" OR "quantitative study" OR "international treaties" OR "international declarations and statements" OR statutes OR laws OR regulations OR resolutions OR codes OR rules OR normativity OR statement | 1,414,423 |
|  | #1 AND #2 | ("research integrity"[Title/Abstract] OR "publication ethics"[Title/Abstract] OR "scientific integrity"[Title/Abstract] OR "research collaboration"[Title/Abstract]) AND (strategies[Title/Abstract] OR policies[Title/Abstract] OR guidelines[Title/Abstract] OR "qualitative study"[Title/Abstract] OR "quantitative study"[Title/Abstract] OR "international treaties"[Title/Abstract] OR "international declarations and statements"[Title/Abstract] OR statutes[Title/Abstract] OR laws[Title/Abstract] OR regulations[Title/Abstract] OR resolutions[Title/Abstract] OR codes[Title/Abstract] OR rules[Title/Abstract] OR normativity[Title/Abstract] OR statement[Title/Abstract]) | 801 |

### 6.2 Web of Science

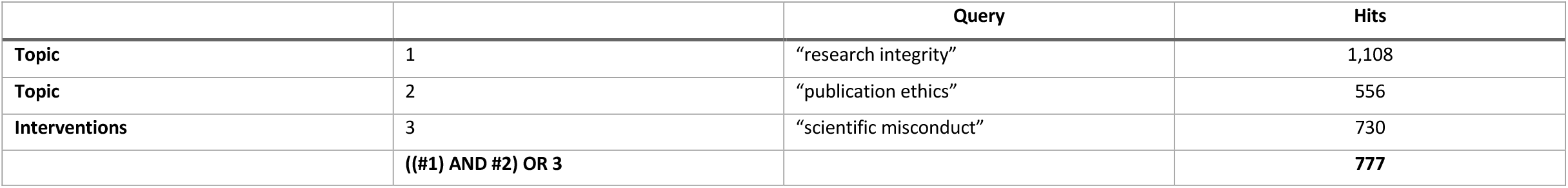

### 6.3 JSTOR

|  |  | Query | Hits |
| --- | --- | --- | --- |
| Topic | 1 | "research integrity" OR "publication ethics" | 580 |
| Population | 2 | researchers OR universities | 1,246,179 |
| Interventions | 3 | "supervision and mentoring" | 38 |
| Type of document | 4 | guidelines OR normativity | 186,518 |
| #1 AND #2 |  | ((("research integrity" OR "publication ethics") AND (researchers OR universities)) | 318 |
| #1 AND #4 |  | ((("research integrity" OR "publication ethics") AND (guidelines OR normativity)) | 246 |

### 6.4 LILACS (September 23^TH^ 2021)

|  |  | Query | HITS |
| --- | --- | --- | --- |
| Topic | 1 | ("Integridad científica") OR ("Integridad en la investigación") OR ("Mala conducta científica") | 380 |
| Interventions | 3 | ("Prácticas de investigación") OR ("Capacitación en investigación") OR ("Tutoría") OR ("Gestión de datos") OR ("Prácticas de publicación") OR ("Autoría") OR ("Ética en las publicaciones") | 806 |
|  | #1 AND # 3 | ((("Integridad científica") OR ("Integridad en la investigación") OR ("Mala conducta científica")) AND ((("Prácticas de investigación") OR ("Capacitación en investigación") OR ("Tutoría") OR ("Gestión de datos") OR ("Prácticas de publicación") OR ("Autoría") OR ("Ética en las publicaciones")) | 556 |

### 6.5 Scopus (September 25 2021)

|  |  | Query | Hits |
| --- | --- | --- | --- |
| Topic | 1 | "research integrity" OR "publication ethics" OR "scientific integrity" OR "research collaboration" | 9012 |
| Type of document | 2 | strategies OR policies OR guidelines OR "qualitative study" OR "quantitative study" OR "international treaties" OR "international declarations and statements" OR statutes OR laws OR regulations OR resolutions OR codes OR rules OR normativity OR statement | 11,376,587 |
|  | #1 AND #2 | ( TITLE-ABS-KEY ( "research integrity" OR "publication ethics" OR "scientific integrity" OR "research collaboration" ) AND TITLE-ABS-KEY ( strategies OR policies OR guidelines OR "qualitative study" OR "quantitative study" OR "international treaties" OR "international declarations and statements" OR statutes OR laws OR regulations OR resolutions OR codes OR rules OR normativity OR statement ) ) | 3,554 |

### 6.6 OECD ILibrary

|  |  | Query | Hits |
| --- | --- | --- | --- |
| Topic | 1 | "research integrity" OR "publication ethics" | 30 |
| Population | 2 | researchers OR universities | 60,544 |
| Interventions | 3 | "supervision and mentoring" | 0 |
| Type of document | 4 | guidelines OR normativity | 28,569 |
| #1 AND #2 |  | (( "research integrity" OR "publication ethics" ) AND ( researchers OR universities )) | 30 |
| #1 AND #4 |  | (( "research integrity" OR "publication ethics" ) AND ( guidelines OR normativity )) | 27 |

